# Profiling Genome-Wide DNA Methylation Patterns in Human Aortic and Mitral Valves

**DOI:** 10.1101/2020.09.10.20190546

**Authors:** Sarah Halawa, Najma Latif, Yuan-Tsan Tseng, Ayman M. Ibrahim, Adrian H. Chester, Ahmed Moustafa, Yasmine Aguib, Magdi Yacoub

## Abstract

Cardiac valves exhibit highly complex structures and specialized functions that include dynamic interactions between cells, extracellular matrix (ECM) and their hemodynamic environment. Valvular gene expression is tightly regulated by a variety of mechanisms including epigenetic factors such as histone modifications, RNA-based mechanisms and DNA methylation. To date, methylation fingerprints of non-diseased human aortic and mitral valves have not been studied. In this work we analyzed the differential methylation profiles of 12 non-diseased aortic and mitral valve tissue samples (in matched pairs). Analysis of methylation data (reduced representation bisulfite sequencing (RRBS)) of 1601 promoters genome-wide revealed 584 differentially methylated (DM) promoters, of which 13 were reported in endothelial mesenchymal trans-differentiation (EMT), 37 in aortic and mitral valve disease and 7 in ECM remodeling. Both functional classification as well as network analysis showed that the genes associated with the DM promoters were enriched for WNT-, Cadherin-, Endothelin-, PDGF- and VEGF-signaling implicated in valvular physiology and pathophysiology. Additional enrichment was detected for TGFB-, NOTCH- and Integrin-signaling involved in EMT as well as ECM remodeling. This data provides the first insight into differential regulation of human aortic and mitral valve tissue and identifies candidate genes linked to DM promoters. Our work will improve the understanding of valve biology, valve tissue engineering approaches and contributes to the identification of relevant drug targets.

## 1. Introduction

Heart valves perform a range of sophisticated functions that ensure unidirectional blood flow during systole, prevent backflow during diastole, enhance coronary blood flow and maintain left ventricular as well as myocardial function [1]. These functions are sustained throughout the human’s lifetime and require tight regulation of the valve cells and extracellular matrix (ECM), which continuously interact together enabling the valves to actively adapt to their complex hemodynamic and biomechanical environments [1,2].

Valves have a unique trilamellar structure. This structure is formed through complex developmental processes starting with the generation of primordial cardiac cushions via endothelial mesenchymal trans-differentiation (EMT) [3]. Remodeling events that include cell differentiation, apoptosis, and ECM remodeling transform these cushions into valve leaflets [3]. Ultimately, the mature valves are composed of a valve endothelial cell (VEC) layer covering a central layered ECM, which comprises collagen, elastin, glycosaminoglycans and is populated by valve interstitial cells (VICs) [4]. The specific diversity and nature of the cells and ECM in each valve-type requires a distinct set of genes to be expressed, which is achieved via molecular regulation by genetic and epigenetic factors [5,6].

Epigenetics refers to heritable phenotype changes that do not involve changes in the DNA sequence itself and includes mechanisms such as histone modifications, RNA-based mechanisms and DNA methylation [7]. DNA methylation is a process by which a methyl group is added to the 5’ carbon of cytosine, which alters the structure of the DNA molecule thus allowing differential regulation of gene expression either through obstructing transcription factor (TF) binding or through the recruitment of methyl-binding proteins, which bind complexes responsible for chromatin remodeling [8]. This process is heritable as well as tissue-specific and plays a major role in various physiological processes linked to cardiogenesis such as cardiomyocyte development, maturation and cardiac regeneration [9,10]. Aberrant changes in methylation profiles are associated with cardiovascular diseases (CVDs) such as ventricular septal defects, tetralogy of Fallot, atherosclerosis as well as other CVDs that lead to end-stage heart failure [11–14].

So far, few studies have addressed how DNA methylation affects valvular disease processes. These studies investigated DNA methylation mechanisms that can transform VICs of stenotic aortic valves into leukotriene-producing immune-like cells via targeted promoter methylation measurement of 5-lipoxygenase (5-LO) [15], and those that lead to the disruption of both the organization of the ECM and the communication between the cells and the ECM in bicuspid aortic valve (BAV) by investigating miR-29 expression level utilizing qRT-PCR [16]. Other studies focused on uncovering associations between methylation changes and the development of rheumatic heart valve disease using ELISA [17], and on detecting the genome-wide DNA methylation landscape underpinning BAV and aortic dissection (AD) via methylation array [18].

To date, DNA methylation in human non-diseased mitral and aortic valves have not been reported. Understanding the unique DNA methylation patterns of non-diseased valves and the methylation differences between different types of human valves is crucial to understanding their biology and susceptibility to disease.

In this study, we investigated the differential methylation fingerprints of 12 human non-diseased aortic and mitral valve samples (in matched pairs) by the assessment of their methylation profiles generated via reduced representation bisulfite sequencing (RRBS). We assessed genes, whose promoters were significantly differentially methylated (DM) on a genome-wide level, and grouped those genes according to their molecular functions, biological processes, protein classes, and pathways. To gain a systems-level understanding of the patterns of methylation of aortic and mitral valve tissue we performed a network analysis, followed by a functional enrichment analysis on the constructed networks to identify significantly DM Gene Ontology (GO) terms and pathways characteristic of these valves. This work provides an initial insight into differential methylation profiles of non-diseased human aortic and mitral valve tissue, and shows the importance of a deep understanding of the tissue and cell-specific mechanisms in the non-diseased status.

## 2. Materials and Methods

### 2.1. Ethics statement and study cohort/samples

This study was approved by the Royal Brompton hospital ethics review board / Brompton and Harefield trust ethics committee (REC approval 10/H0724/18) and is abiding by all the standards of the Declaration of Helsinki. Written informed consent was obtained from the donors prior to their inclusion in the study. Twelve non-diseased valves free from calcification (6 aortic and 6 mitral valves; 10 males: 2 females; age range 42 – 64 years, mean age 52.2 years, SD 9.9682) were used in this study. After applying inclusion/exclusion criteria three of the twelve valves were excluded from the downstream analysis (**Figure S1**). The non-diseased valves were obtained from unused valves of healthy donor hearts, who died of non-cardiac diseases (**Table 1**). History, macroscopic, and microscopic evaluation were additionally performed to make sure that the donor hearts chosen are free from cardiovascular and valvular complications. The exclusion criteria of donor hearts were previously described in [19].

**Table 1.** Cohort demographic and clinical characteristics.

| Donor No. | Gender | Age | Cause of Death |
| --- | --- | --- | --- |
| 1. | Male | 44 | Transplant recipient |
| 2. | Male | 57 | Intracerebral hemorrhage |
| 3. | Male | 42 | Intracranial thrombosis |
| 4. | Male | 44 | Transplant recipient |
| 5. | Male | 62 | Intracerebral hemorrhage |
| 6. | Female | 64 | Intracerebral hemorrhage |

### 2.2. Tissue Sampling and DNA Extraction

Aortic and mitral valves from donor hearts were provided by the Royal Brompton Valve biobank. The ischemic time for fresh tissue harvest was set to not exceed 24 h. The aortic and mitral valve leaflets were excised and separately deendothelialized using collagenase II for 10 minutes at 37 °C. The leaflets were then washed using PBS, snap frozen and stored in −80 °C for DNA extraction. DNA was isolated from deendothelialized tissue using the FitAmp™ Blood and Cultured Cell DNA Extraction Kit (Epigentek, NY, USA, catalog #: P-1018) and was subsequently eluted in TE buffer in a total volume of 40 μl. DNA was finally quantified and quality controlled via fluorescence.

### 2.3. Bisulfite Conversion, Library Preparation and Sequencing

For each sample, 300 ng of DNA was digested for 2 hours with the MSP1 enzyme (20U/sample at 37°C) followed by 2 hours with TaqαI (20U/sample at 65°C). Digested, CGI enriched DNA fragments less than 300 bps in length, were selected for and subsequently collected for bisulfite treatment. Bisulfite treatment was performed using the Methylamp DNA Bisulfite Conversion Kit (Epigentek, NY, USA, catalog #: P-1001). Bisulfite conversion efficiency of the bisulfite-treated DNA was determined by RT-PCR using two pairs of primers against bisulfite-converted DNA (b-actin) and against unconverted DNA (GAPDH), for the same bisulfite-treated DNA samples. Conversion was deemed successful, if more than 99% of the DNA were converted (Epigentek, NY, USA).

For library preparation, first DNA end polishing and adaptor ligation was performed. This was followed by library amplification using indexed primers and library purification. The final purified library was eluted in 12 μl of water. Assessment of library quality was done via bioanalyzer and KAPA library quantification (Roche, CA, USA). Finally, 10 nM of sample libraries were subjected to single-end enhanced RRBS on Illumina HiSeq 2500.

### 2.4. Data Quality Control (QC) and Processing

A summary of the bioinformatics analysis workflow can be found in **Figure 1**. First, raw reads were subjected to QC using FastQC version 0.10.1 [20]. Trim Galore version 0.3.7 was then used to remove low quality reads, adapters as well as RRBS-related residues that are artificially added during the end-repair step [21]. Trimmed reads were then mapped to the UCSC Homo sapiens genome sequence (version hg19) using Bismark version 0.13.0 [22]. To permit only up to one mismatch per seed region, the option “-n 1” was set for Bowtie version 1.0.0 utilized by Bismark [23]. Samtools version 0.1.19-96b5f2294a, was utilized to sort the SAM file storing bisulfite alignment and methylation calling data produced by Bismark and to remove duplicate reads that result from PCR amplification [24]. Methylation information was extracted from Bismark’s sorted and filtered mapping results at base resolution using Bismark’s methylation extraction software [22]. The subsequent analysis was performed in the CpG context. The R package methylKit version 0.9.2 was used for further analysis of the Bismark methylation extraction reports [25]. Samples that generated a minimum of 60 million reads (**Figure S1**) were included in the analysis and processed methylKit. Methylation information found in the aforementioned extraction reports was summarized by methylKit over RefSeq promoters, defined as regions located 1 kb before or after a transcription start site [26]. Coverage for each promoter was calculated as the sum of the methylated and unmethylated cytosines. Percentage of methylation for each promoter was calculated as the number of methylated cytosines divided by the coverage multiplied by 100. The promoters were subsequently filtered based on coverage (minimal 5 and maximal 99.9 percentile) and merged for comparative analysis with only those promoters that are covered in all replicates being considered. Additional QC steps can be found in the Supplementary Materials Online (**Figure S2, Figure S3**). RRBS data was deposited in EMBL-EBI’s European Genome-phenome Archive (EGA) and is accessible through EGA’s accession number EGAD00001006303.

**Figure 1.**
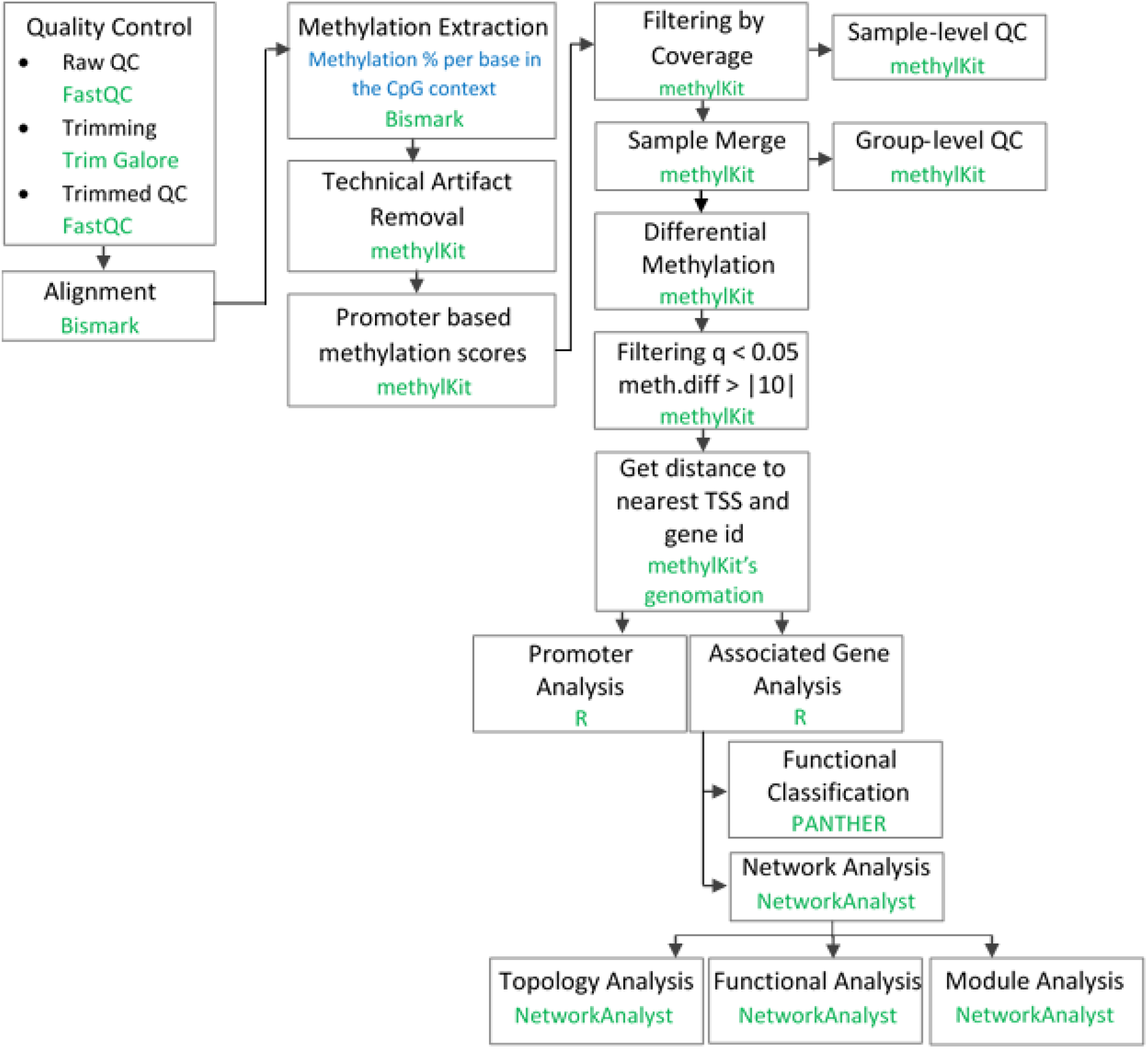
Summary of the data analysis workflow.

### 2.5. Differentially Methylated Region (DMR) Analysis

The difference in the methylation level of a promoter between aortic and mitral tissue was calculated as the difference between the weighted average of the percent methylation values of all mitral tissue samples and the weighted average of the percent methylation values of all aortic tissue samples at this promoter. A positive methylation difference (hypermethylation) indicated increased methylation in the promoter associated with mitral compared to aortic tissue. The significance of this difference was evaluated by methylKit using logistic regression [25]. To correct for multiple hypothesis testing, the sliding linear model (SLIM) method was used by methylKit [25]. DM promoters were finally filtered with the cut-off chosen as a methylation difference that is larger than 10 % and a q-value that is less than 0.05. Annotation of DMRs with genic features, primarily promoters, was carried out by methylKit using the genomation package [25].

### 2.6. Pathway Enrichment Analysis

The Protein Analysis Through Evolutionary Relationships (PANTHER) classification system was utilized to categorize genes whose promoters were found to be DM according to their molecular functions, biological processes, protein classes, pathways and cellular components [27].

### 2.7. Construction of Protein-Protein Interaction (PPI) Networks

Genes associated with DM promoters were used as seed genes to construct PPI networks using NetworkAnalyst [28]. The International Molecular Exchange (IMEx) Interactome database, which contains literature-curated comprehensive data from InnateDB, was used by NetworkAnalyst for the generation of the generic network [28]. For network construction, methylation difference values of alternative promoters of the same seed gene were replaced by their average before proceeding to network construction [28]. The resulting network was trimmed to the minimum to encompass only nodes that connect the original seed genes. Functional enrichment analysis of all seed genes was performed using NetworkAnalyst’s Function Explorer using GO, KEGG and Reactome databases. Enrichment p-values were computed based on the hypergeometric test utilized by the Function Explorer [28]. NetworkAnalyst’s Module Explorer, was used to identify smaller significantly densely connected subnetworks using the Walktrap algorithm [28]. Visualization of PANTHER, KEGG and Reactome pathway terms was performed using the GOplot R package [29].

## 3. Results

### 3.1. Aortic and mitral valves show different methylation signatures in 584 promoters

DMR analysis identified 584 significantly DM promoters, of which 305 showed increased methylation in mitral and 279 in aortic valve tissue (**Figure 2, Figure S4, Table S1**). The gene Repulsive Guidance Molecule A (RGMA) was associated with the most significantly DM promoter followed by TBC1 Domain Family Member 32 (TBC1D32), B-Cell Lymphoma 3 (BCL3) and the long non-coding RNA RP11-1149O23.3 among others (**Figure 2, Table S1**).

**Figure 2.**
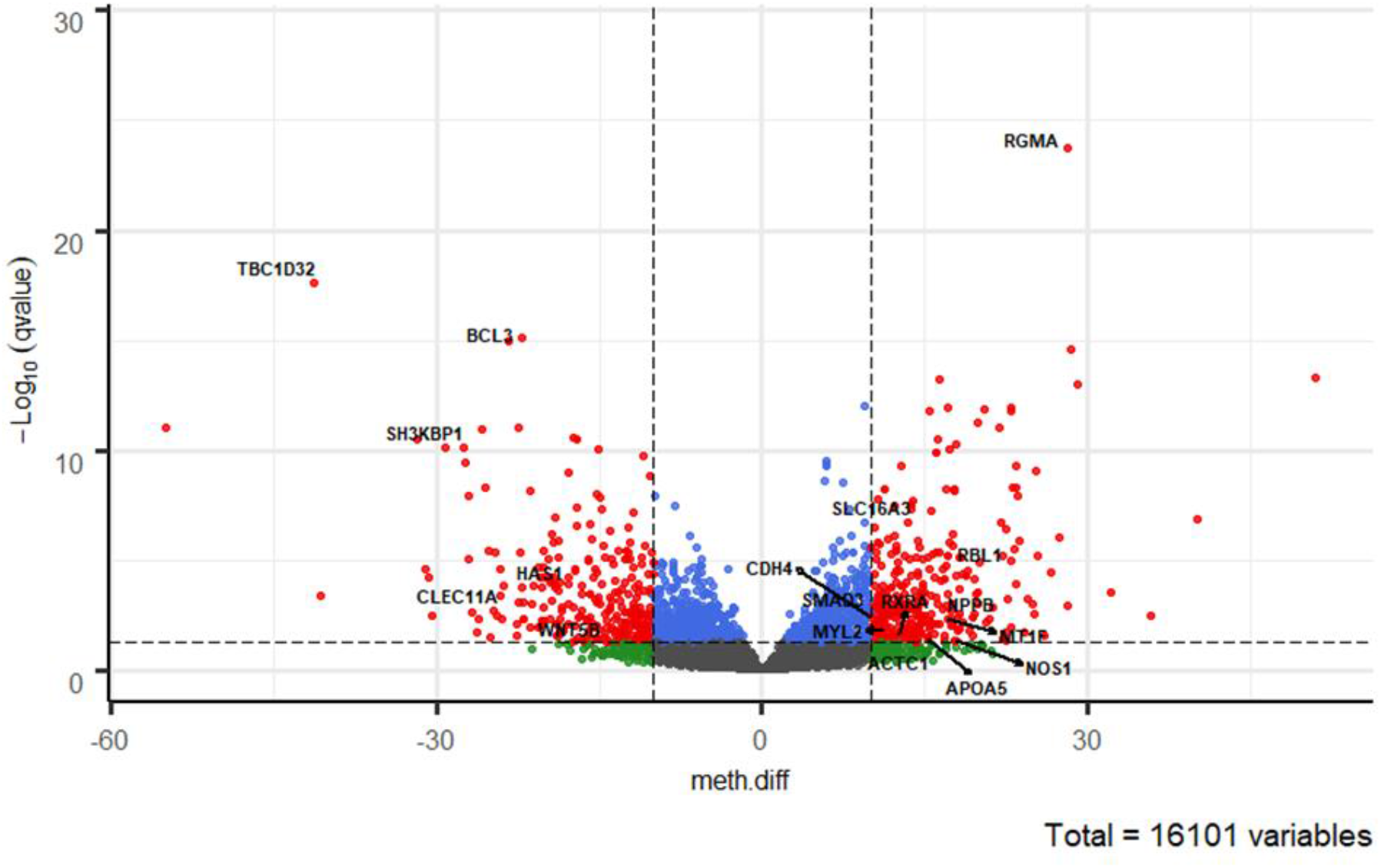
Volcano plot of promoter methylation profiles. The vertical lines x = −10 and x = 10 represent our chosen methylation difference (meth.diff) cut-off value of |10|. meth.diff is calculated as described in the Methods. The horizontal line (y = −log10(0.05) = 1.3) shows our chosen q-value cut-off of 0.05. The circles represent the 1601 promoters, of which 584 are differentially methylated (DM). Each circle in the volcano plot represents a promoter with its meth.diff and −log10(q-value). Grey circles denote promoters that are not DM. Green circles represent promoters that are biologically, but not statistically significant. Blue circles depict the opposite trend. Red circles show promoters that are both statistically and biologically significant. Genes linked to DM promoters discussed in this manuscript are indicated on the plot.

We then functionally classified all genes associated with DM promoters according to pathways and functional categories (**Figure 3 a-e, Figure S5**) and identified their methylation direction in (**Table S1**). Key valve-related pathways such as Wingless/Integrated (WNT)-, Cadherin-, Transforming Growth Factor Beta (TGFB)-, Integrin-, Endothelin-, Platelet-Derived Growth Factor (PDGF)-, NOTCH signaling as well as angiogenesis and general transcription regulation were identified utilizing the PANTHER Pathway database (**Figure 3 a, Table S2**).

**Figure 3.**
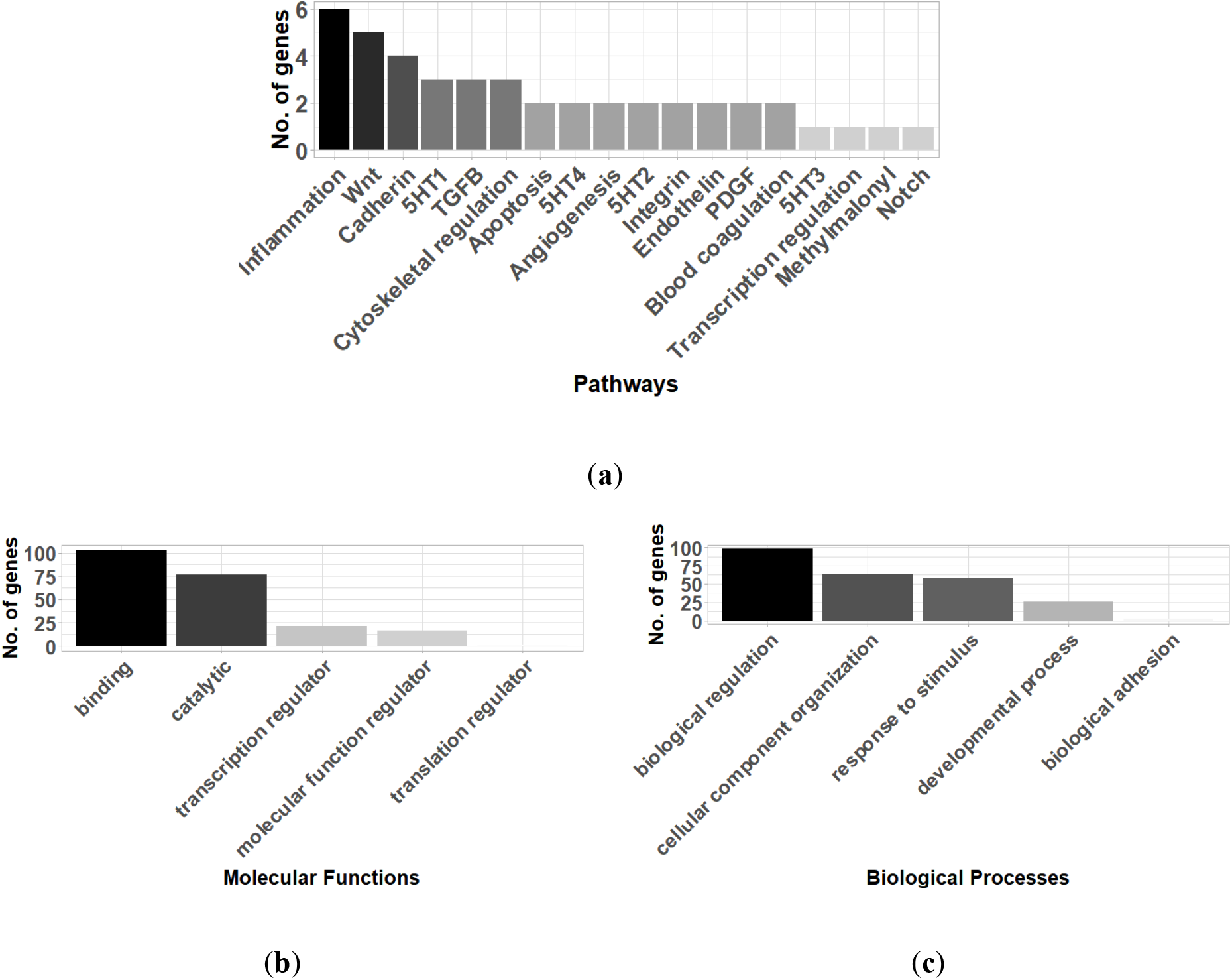
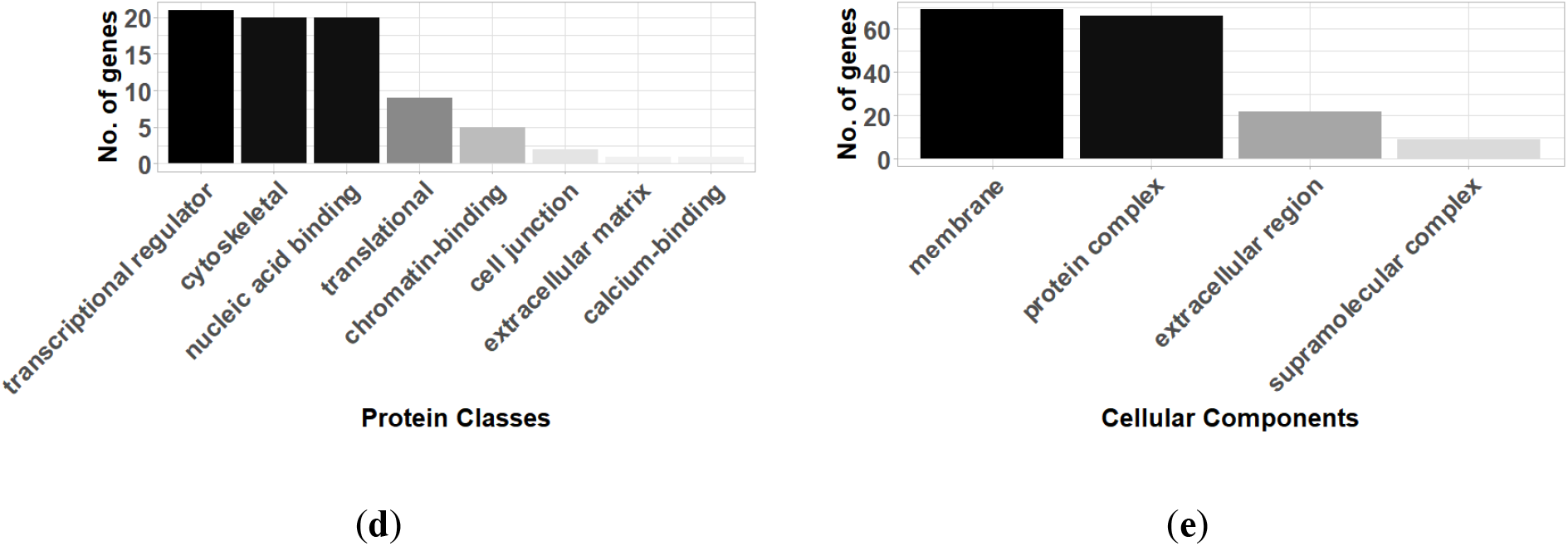
Functional classification of genes associated with differentially methylated promoters (q-value < 0.05 & meth.diff > |10|) between aortic and mitral tissue according to (**a**) Pathways, (**b**) Molecular Functions, (**c**) Biological Processes, (**d**) Protein Classes and (**e**) Cellular Components (selected results, the remaining can be found in **Figure S5**– analysis performed using PANTHER).

When the genes were classified by their molecular function, 69.5% of the genes had a binding and catalytic activity including key genes such as Nitric Oxide Synthase 1 (NOS1) (q-value = 0.04, meth.diff = 17.9 %). Other genes exhibited transcription regulator-, translation regulator- and molecular function regulator activities. The latter category included relevant genes such as Apolipoprotein A5 (APOA5) (q-value = 0.037, meth.diff = 15.3%) and Natriuretic Peptide B (NPPB) (q-value = 0.0027, meth.diff = 15.58%) (**Figure 3b, Table S3**).

Among the relevant biological processes categorizing the genes were “biological regulation” and “cellular component organization or biogenesis”. The former included notable genes such as Metallothionein 1F (MT1F) (q-value = 0.004, meth.diff = 17.07%) and the latter included two pertinent ones namely Hyaluronan Synthase 1 (HAS1) (q-value = 0.00015, meth.diff = 22.15%) as well as Actin Alpha Cardiac Muscle 1 (ACTC1) (q-value = 0.049, meth.diff = 11.23%) (**Figure 3c, Table S4**).

Genes were also grouped into relevant protein classes such as gene-specific transcriptional regulator proteins, which included SMAD Family Member 3 (SMAD3) (q-value = 0.0019, meth.diff = 10.25%), cytoskeletal proteins, which contained Myosin Light Chain 2 (MYL2) (q-value = 0.013, meth.diff = 11.14%) and chromatin-binding protein, which encompassed RB Transcriptional Corepressor Like 1 (RBL1) (q-value = 9.5e-06, meth.diff = 22.98%) (**Figure 3d, Table S5**).

Finally, the cellular component categories into which the genes could be classified were membrane-and extracellular regions among others. Relevant genes such as C-Type Lectin Domain Containing 11A (CLEC11A) (q-value = 0.002, meth.diff = 24.72%) were included in the latter and Solute Carrier Family 16 Member 3 (SLC16A3) (q-value = 1.78e-07, meth.diff = 13.4%) in the former category (**Figure 3e, Table S6**).

### 3.2. Network analysis enabled a systems-based assessment and revealed additional valve-related pathways such as Hypoxia-Inducible Factor 1 (HIF-1) and Vascular Endothelial Growth Factor (VEGF) signaling

After functionally categorizing the genes, whose promoters were significantly DM between the aortic and mitral valves on a genome-wide level, we wanted to assess whether we can predict significantly enriched PPIs among those genes as well as the proteins that are necessary to interconnect them to enable a systems-level analysis.

For that purpose, we mapped those DM genes onto a generic PPI database (see Methods) and constructed a PPI network, which ultimately comprised 715 nodes (genes) of which 308 were seed genes and 2131 edges (PPIs) (**Figure 4a, Table S7**). Among the network’s constituent proteins were several hub nodes, the most connected of which were Ubiquitin C (UBC) (Betweenness centrality = 129377.98; Degree = 200), SMAD3 (Betweenness centrality = 27152.04; Degree = 75), Ubiquitin Like 4A (UBL4A) (Betweenness centrality = 13019.92; Degree = 50), Ribosomal Protein S3 (RPS3) (Betweenness centrality = 9508.1; Degree = 50), Retinoid X Receptor Alpha (RXRA) (Betweenness centrality = 12661.72; Degree = 47) and SH3 Domain Containing Kinase Binding Protein 1 (SH3KBP1) (Betweenness centrality = 13005.65; Degree = 44) (**Figure 4a, Table S7**).

**Figure 4.**
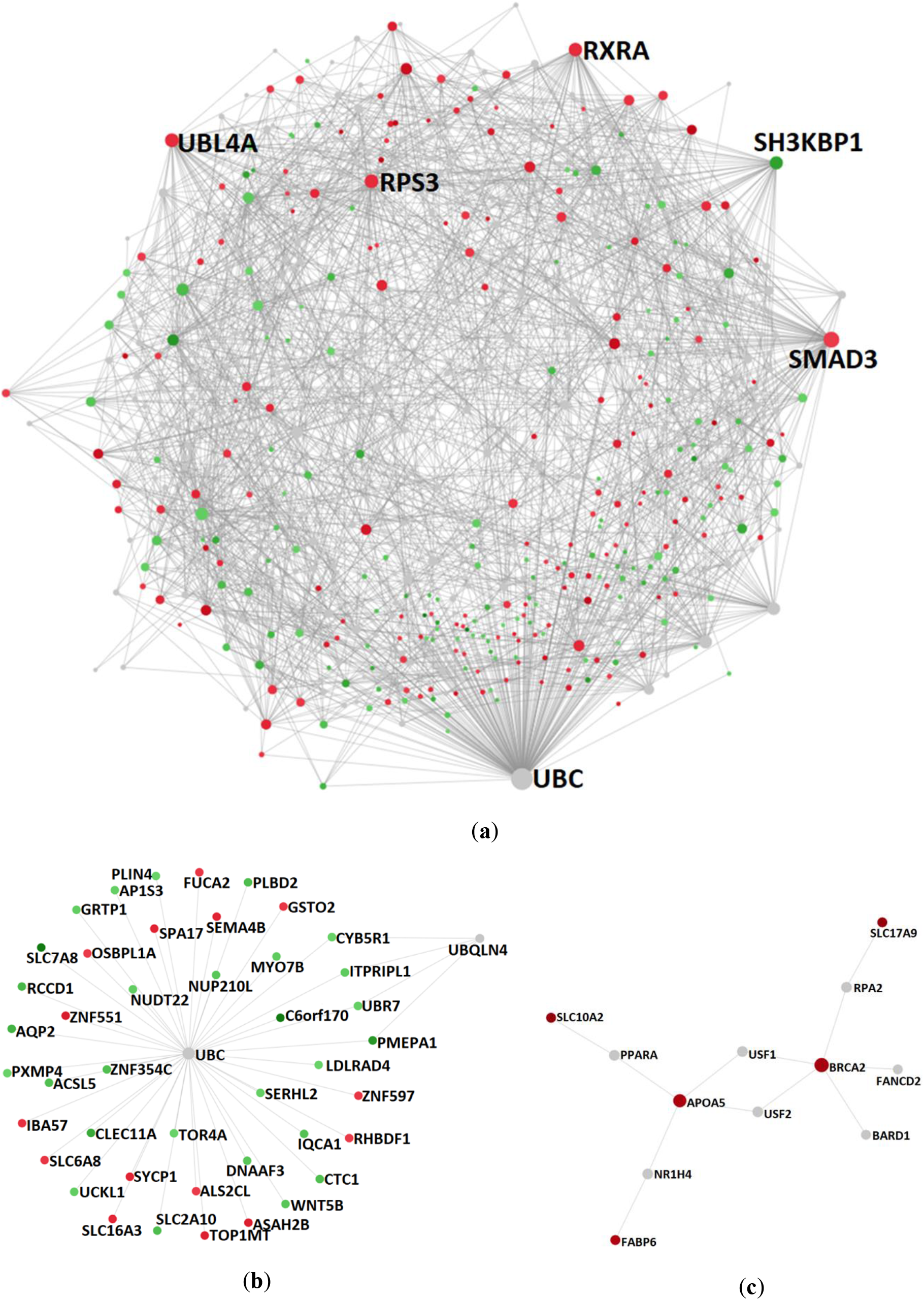
PPI networks constructed upon the genes associated with differentially methylated promoters between non-diseased aortic and mitral valve tissue and the proteins that are necessary to interconnect them: (**a**) complete network, (**b**) subnetwork 1 (p-value 3.04e-13) and (**c**) subnetwork 2 (p-value 0.047). Depicted nodes are classified according to their methylation direction: nodes representing genes showing increased methylation in mitral compared to aortic tissue (red), nodes depicting increased methylation in aortic versus mitral tissue (green) and nodes representing genes that are not part of the input dataset (grey). Node sizes are proportional to their betweenness centrality values. Betweenness centrality reflects the number of shortest paths passing through a node, while degree refers to the number of connections/edges/PPIs that a node has to other nodes. Hub nodes (**a**) UBC, SMAD3, UBL4A, RPS3, RXRA, (**b**) UBC and (**c**) BRCA2 and APOA5 can be clearly identified on the respective networks.

In addition to inspecting the topological properties of the constructed network, we performed a functional enrichment analysis of both the hyper- and hypomethylated proteins of the subnetwork as well as the proteins connecting them to investigate, whether we can detect relevant significantly enriched pathways and functions.

Many pathways, which are pertinent to valvular mechanisms, were identified via KEGG pathway enrichment analysis including apoptosis, Nuclear Factor Kappa-light-chain-enhancer of activated B cells (NF-Κb) signaling pathway, fluid shear stress and atherosclerosis, Tumor necrosis factor (TNF) signaling pathway, osteoclast differentiation, Interleukin 17 (IL-17) signaling pathway, HIF-1 signaling pathway, regulation of actin cytoskeleton, VEGF signaling pathway and TGFB signaling pathway (FDR <0.05, **Table 2**).

**Table 2.**
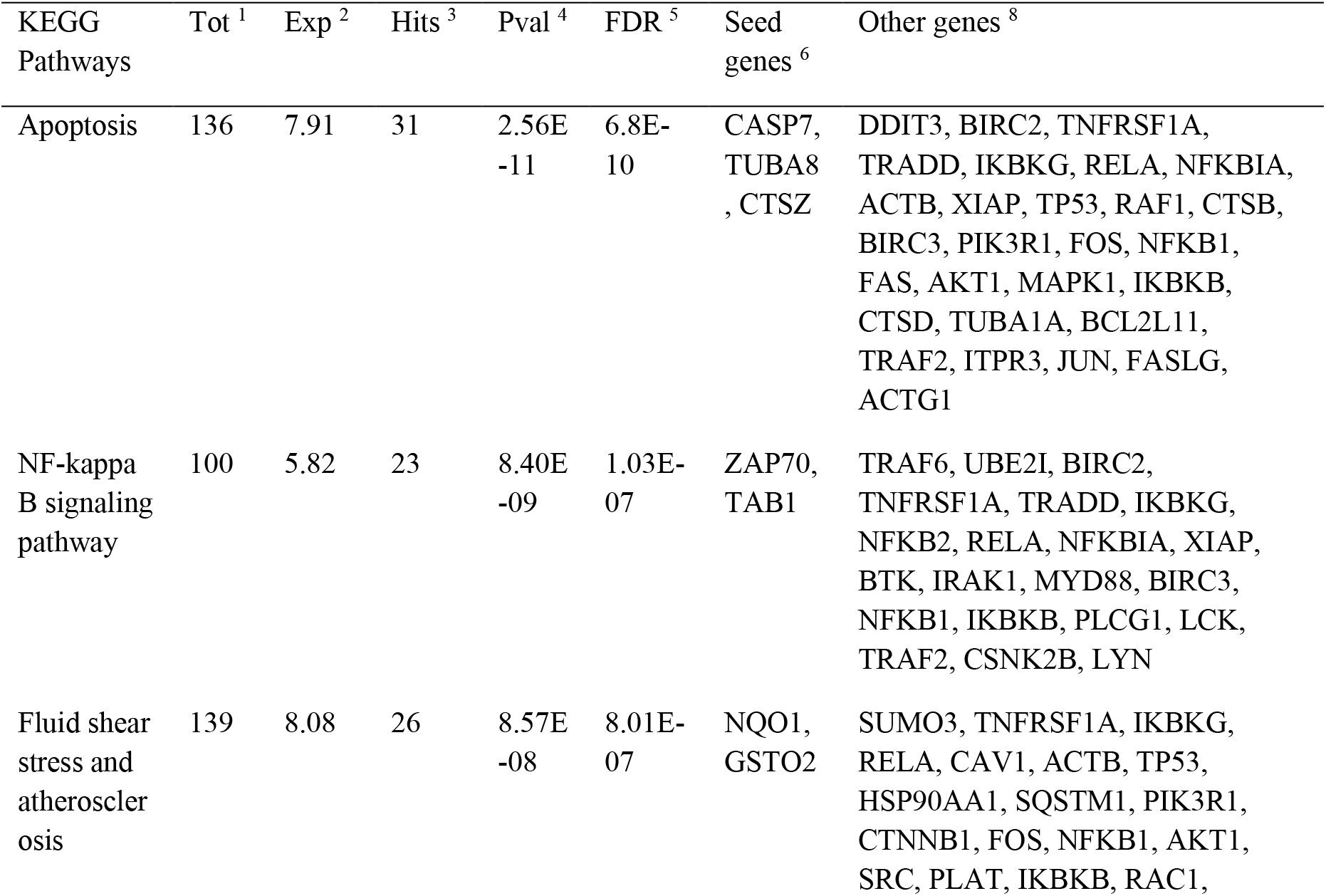
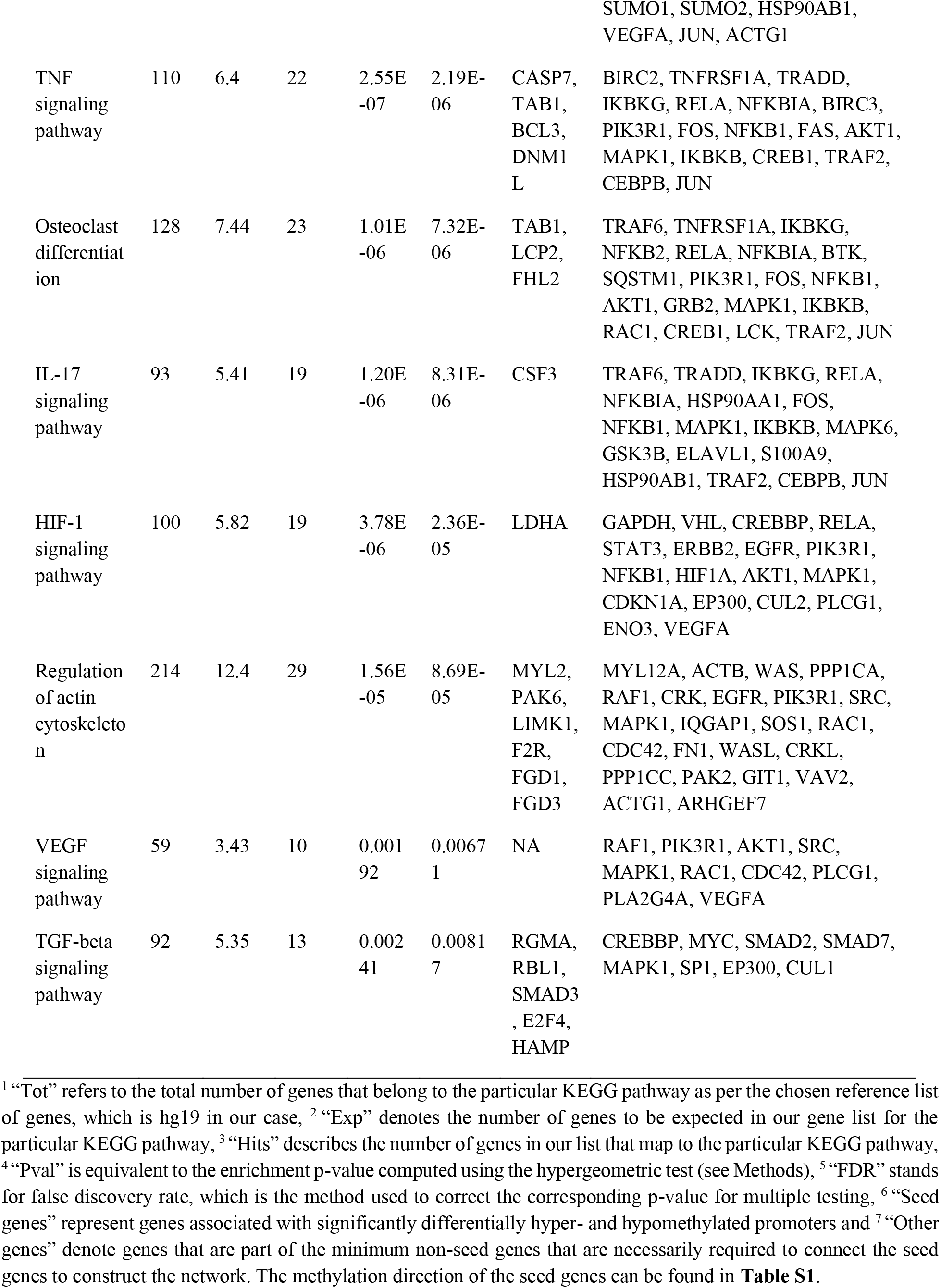
List of pathways resulting from enrichment analysis of the network (**Figure 4a**) constructed upon the genes associated with the differentially methylated hypo- and hypermethylated promoters using the KEGG pathway database.

To investigate whether relevant significantly enriched pathways are present independent of the choice of the pathway database, we furthermore performed Reactome enrichment analysis, which exclusively highlighted additional key valve-related pathways such as immune response, regulation of lipid metabolism, PDGF, NOTCH1, Fibroblast Growth Factor Receptor (FGFR) and transcription (FDR <0.05, **Table S8**). Similar to KEGG, it showed enrichment for apoptosis-, TFGB-, interleukin- and hypoxia-related pathways (**Table 2, Table S8**).

After inspecting significant valve-related pathways, we checked whether there are enriched biological processes, molecular functions and cellular components related to the molecular regulation of valve-related processes based on GO databases. Relevant biological processes included transcription, apoptosis, growth factor signaling as well as regulation of cellular component organization (FDR <0.05, **Table S9**). Molecular functions associated with transcription as well as histone acyltransferase activity such as chromatin-, histone deacetylase-, TF-, SMAD- and NF-κB-binding were among the pertinent significantly enriched molecular functions (FDR <0.05, **Table S10**). Finally, the most relevant significantly enriched cellular components were Transcription Factor II D (TFIID) complex, spliceosomal complex, chromatin, histone deacetylase complex as well as actin cytoskeleton (FDR <0.05, **Table S11**).

In addition to examining the network structure as a whole, we performed a module analysis (see Methods), to identify subnetworks that show a significantly increased connection density compared to other modules of the parent network. The first identified subnetwork (p-value 3.04e-13) comprised 45 nodes, of which 43 were seed genes and contained the following hub nodes: UBC, also identified as a hub protein of the parent network (Betweenness centrality = 43; Degree = 939) and Ubiquilin 4 (UBQLN4), not identified previously (Betweenness centrality = 3; Degree = 4) (**Figure 4 b, Table S12, Table S13**).

The second subnetwork (p-value 0.047) contained 12 nodes, of which 5 were seed genes with Breast And Ovarian Cancer Susceptibility Protein 2 (BRCA2) (Betweenness centrality = 33.5; Degree = 5) as well as APOA5 (Betweenness centrality = 32.5; Degree = 4) being its main hub proteins. Interestingly, all of the module’s constituent seed nodes exhibited increased methylation in mitral compared to aortic tissue (**Figure 4c, Table S14**). Relevant enriched pathways in this module included homologous recombination (KEGG/Reactome) and Peroxisome Proliferator-Activated Receptor (PPAR) signaling pathway (KEGG) (FDR <0.05, **Table S15**). The module’s biological processes contained lipid homeostasis and cellular response to external stimulus (FDR <0.05, **Table S15**). Finally, the most relevant enriched molecular functions encompassed DNA-, enzyme- and TF-binding (FDR <0.05, **Table S15**).

### 3.3. Global analysis of detected pathways and genes shows the role of methylation in EMT and ECM remodeling

All the utilized analysis methods of genes and pathways show that DNA methylation plays a crucial role in valve development and disease. Indeed, some of the detected genes and pathways are involved in developmental processes such as cardiogenesis, EMT and protein QC and/or diseases such as myxomatous mitral valve (MMV), BAV and AD (**Table 3,Table 4**).

**Table 3.** Genes associated with differentially methylated promoters between aortic and mitral valve tissue and their involvement in valve development and disease.

| Gene | Description | Development | Disease |
| --- | --- | --- | --- |
| NOS1 | nitric oxide synthase 1 (neuronal) | heart [46] | BAV <sup>1</sup> [46] |
| ACTC1 | actin, alpha, cardiac muscle 1 | heart [47] | MMV <sup>2</sup> [49] |
| MYL2 | myosin, light chain 2, regulatory, cardiac | heart [48] |  |
| MT1F | metallothionein 1F |  | MMV [50] |
| CLEC11A | C-Type lectin domain containing 11A |  | MMV [51] |
| RBL1 | retinoblastoma-like 1 (p107) |  | BAV [52] |
| SLC16A3 | solute carrier family 16, member 3<br>(monocarboxylate transporter) |  | BAV [53], AS <sup>3</sup> [59] |
| NPPB | natriuretic peptide B | EMT <sup>4</sup> [54] | MR <sup>5</sup> [55] |
| CDH4 | cadherin 4, type 1, R-cadherin (retinal) | valve [56] |  |
| HAS1 | hyaluronan synthase 1 |  | CAVD <sup>6</sup> [57] |
| WNT5B | wingless-type MMTV integration site<br>family, member 5B |  | CAVD [58] |
| Ubiquitin-<br>related<br>genes |  | protein QC <sup>7</sup> in<br>the heart [62] | BAVs [60], atherosclerosis<br>[61] |
| SMAD3 | SMAD family member 3 | cardio-genesis<br>[63] | AD <sup>8</sup> [64] |
| RXRA | retinoid X receptor, alpha |  | Valve malformation [65] |
| SH3KBP1 | SH3-domain kinase binding protein 1 |  | CAVD [66] |
| APOA5 | apolipoprotein A-V |  | AS [67–69] |

<sup>1</sup> BAV: bicuspid aortic valve; <sup>2</sup> MMV: myxomatous mitral valve; <sup>3</sup> AS: aortic valve stenosis; <sup>4</sup> EMT: endothelial mesenchymal trans-differentiation; <sup>5</sup> MR: mitral valve regurgitation; <sup>6</sup> CAVD: calcific aortic valve disease; <sup>7</sup> QC: quality control; <sup>8</sup> AD: thoracic aortic aneurysm and dissection.

**Table 4.** Pathways involving the genes associated with the differentially methylated promoters between aortic and mitral valve tissue and their implication in valve development and disease.

| Pathway | Description | Development | Disease |
| --- | --- | --- | --- |
| TGFB signaling | Transforming growth factor beta signaling | EMT <sup>1</sup> [3] | ECM <sup>2</sup> remodeling [71], MVP <sup>3</sup> [72], AS <sup>4</sup> [73] |
| NOTCH signaling |  | EMT [3], [70] | BAV <sup>5</sup> [70] and AS [70] |
| FGF signaling | Fibroblast growth factor signaling | EMT [74] | Valve malformation [74] |
| WNT signaling |  | heart, EMT [3,58] |  |
| Cadherin signaling |  | EMT [75] |  |
| PDGF signaling | Platelet-derived growth factor | heart [76] |  |
| VEGF signaling |  | EMT [3] |  |
| Integrin signaling |  | EMT [3], cell-ECM [77,78] |  |
| HIF-1 signaling |  |  | RMV <sup>6</sup>, MMV <sup>7</sup> [79] |
| Angiogenesis |  |  | RMV [80] |
| IL-17 signaling |  |  | IE <sup>8</sup> [81] |
| NF-κB signaling |  |  | AS [82] |
| TNF signaling |  |  | AS [83] |
| Osteoclast differentiation pathways |  |  | AS [84] |
| Endothelin signaling |  | VICs <sup>9</sup> regulation by VECs <sup>10</sup> [87] | AS [85] |
| Apoptotic pathways |  |  | AS [86] |
| PPAR signaling |  | lipid metabolism [88] |  |

<sup>1</sup> EMT: endothelial mesenchymal trans-differentiation; <sup>2</sup> ECM: extracellular matrix; <sup>3</sup> MVP: mitral valve prolapse; <sup>4</sup> AS: aortic valve stenosis; <sup>5</sup> BAV: bicuspid aortic valve; <sup>6</sup> RMV: rheumatic mitral valve; <sup>7</sup> MMV: myxomatous mitral valve; <sup>8</sup> IE: infective endocarditis; <sup>9</sup> VICs: valve interstitial cells; <sup>10</sup> VECs: valve endothelial cells.

We further generated a GOChord plot to link the pathways in **Table 4** to their constituent genes, which are associated with DM promoters (**Figure 5a**). The plot revealed several pathways sharing the same genes, such as NF-κB signaling, TNF signaling and osteoclast differentiation, which contain TGF-Beta Activated Kinase 1 Binding Protein 1 (TAB1) and Integrin- and PDGF signaling, which include Collagen Type V Alpha 3 (COL5A3) (**Figure 5 a**).

A GOCircle plot visualizes the significance of each pathway in **Table 4** and to show the direction of promoter methylation of their constituent genes (**Figure 5 b**). The most significant pathways in **Table 4** were NF-κB, TNF signaling and osteoclast differentiation, which share TAB1 (**Figure 5 a**) and are implicated in aortic stenosis (AS) (**Table 4**).

**Figure 5.**
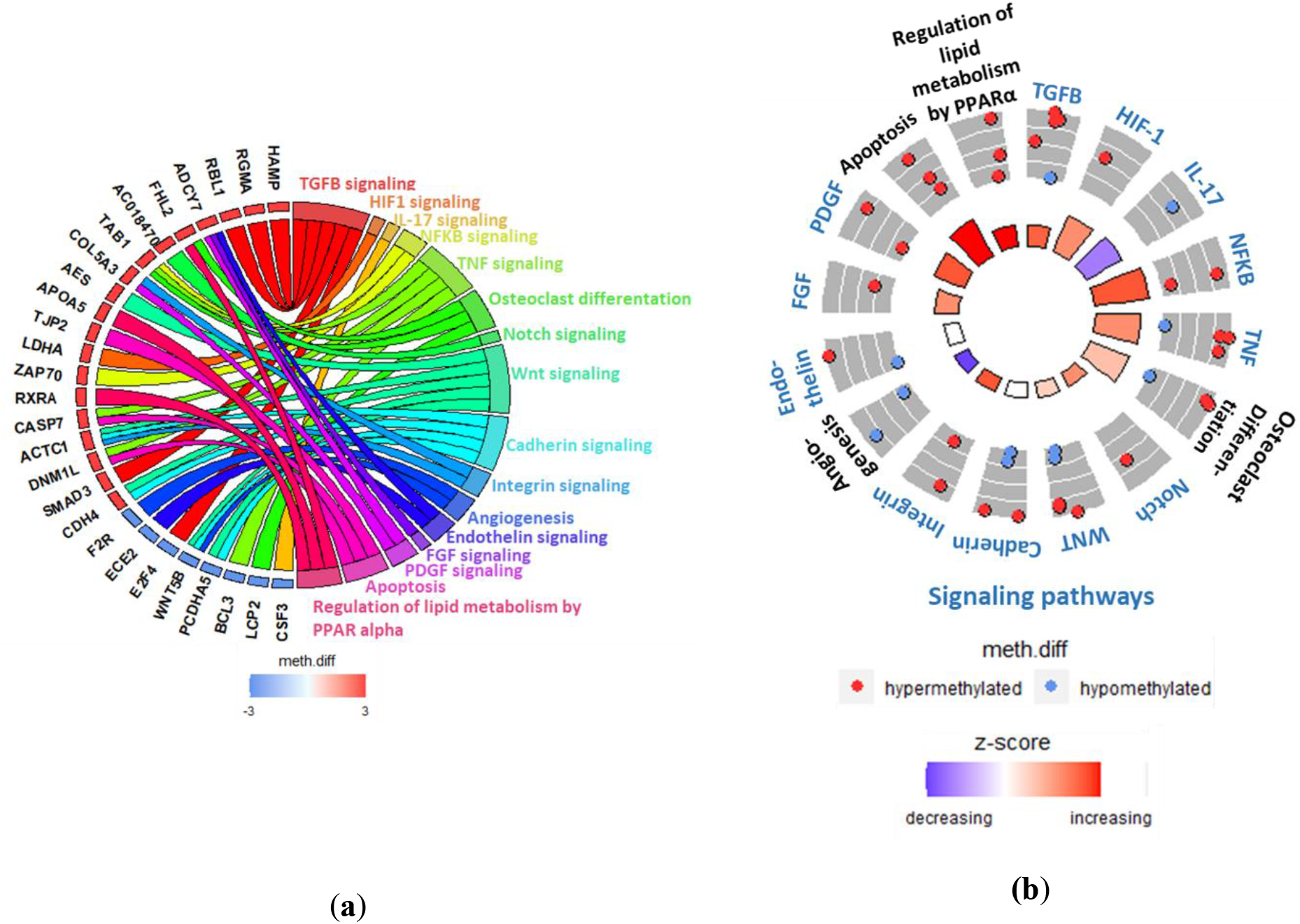
(**a**) GOChord plot linking selected pathways to their constituent genes, which are associated with DM promoters. Blue-to-red colors next to the selected genes reflect their meth.diff values. (**b**) GOCircle plot representing the relevant pathways and their constituent genes that are associated with DM promoters. The inner circle consists of bar plots, whose heights reflect the significance of the pathway and whose color reflect the z-score, which approximates the overall direction of change in methylation for each pathway. The outer circle shows scatterplots of the meth.diff values of each of the pathways’ constituent genes.

## 4. Discussion

This study explores the DNA methylation landscape in human non-diseased heart valves. Heart valves have a complex structure and function that are sensitive to their environment and exhibit characteristic phenotypic and functional differences. Valve-type specific differences begin to appear during valve formation and development and are expressed in the valves’ distinct anatomical structures, environmental milieus and susceptibility to disease. In this work we focus on two of the four valves, the mitral and aortic valves and their epigenetic profiles. On the developmental level, the mature mitral valve is formed from atrioventricular (AV) cushions generated by EMT, while the mature aortic valve is created from cushions deposited at the outflow tract (OFT) [30]. On the anatomical level, the mitral and aortic valves differ in that the mitral valve is composed of two leaflets, the anterior and posterior leaflets, while the aortic valve is composed of three cusps, the left-, right- and non-coronary cusps [31]. Additionally, the mitral valve is anchored by a specialized structure consisting of the chordae tendineae and papillary muscles, whereas the distinct shape of the aortic valve provides a unique self-contained support configuration [32]. The entire aortic valve machinery is dynamic as its components move spatially as well as alter their shape and size in response to different conditions of the cardiac cycle [33]. Due to this dynamism it is described as the “living aortic valve”[34]. On the hemodynamic level, the valves are exposed to different forces with the mitral valve, being exposed to higher pressures than the aortic valve [1]. All the described above makes the mitral and aortic valves susceptible to different diseases with the mitral valve being more prone to mitral valve prolapse (MVP), rheumatic mitral valve (RMV) stenosis, degenerative mitral regurgitation (MR) and MMV disease, whereas the aortic valve being more predisposed to AS, calcific aortic valve disease (CAVD) and BAV disease [35]. To understand the aforementioned differences that exist despite having the same genetic basis, it is important to understand mechanisms of epigenetic regulation of major valve-type specific affected genes and pathways. Our work is focused on profiling genome-wide DNA methylation patterns based on a small cohort of matched non-diseased donor heart valves to enable the characterization of valve-type associated differences as well as their separation from valve disease-related ones. By having matched donors, confounding effects such as gender, age, valve/patient health status are reduced further highlighting the accuracy of the generated aortic and mitral valve methylation profiles.

We used RRBS to measure DNA methylation as it targets CpG-rich islands and promoters genome-wide [36]. It is crucial to couple this sequencing method with suited bioinformatics workflows, that rely on rigorous QC of RRBS-characteristic issues as well as of bisulfite sequencing-specific parameters such as the efficiency of bisulfite conversion [37]. RRBS thus can allow for a comprehensive view not limited by predefined sets of CpG loci probes [38].

In this study, 584 of 1601 promoters were found to be DM between aortic and mitral tissue and their associated genes associated were found to be implicated in valvular health and disease mechanisms. RGMA was associated with the most significantly DM promoter. It is a member the RGM protein family, the first known BMP selective co-receptor family able to induce BMP signaling that is dysregulated in CAVD [39,40]. TBC1D32, the gene associated with the second most DM promoter, is implicated in the pathogenesis of ciliopathies in humans [41], which are caused by defects in the human primary cilium known to play a role in establishing left-right asymmetry during heart development [42], to restrain ECM production during physiological aortic valve development and to play a role in the etiology of BAV in humans [43]. Finally, BCL3, the gene associated with the third most DM promoter, is known to play a role in atherosclerosis [44], with atherosclerosis-like lesions potentially leading to AS [45].

Further genes, that were associated with DM promoters (summarized in **Table 3**) include NOS1, ACTC1 and MYL2, which play key roles in heart development [46–48], with ACTC1 additionally being implicated in MMV and NOS1 in BAV [46,49]. MT1F and CLEC11 further contribute to MMV [50,51], while RBL1 and SLC16A3 to BAV [52,53]. NPPB is involved in EMT by exhibiting excessive synthesis of the cardiac jelly, a precursor of the cushions, in zebrafish and is overexpressed in the ventricles of patients with chronic volume overload caused by regurgitant mitral valve lesions [54,55]. Cadherin-4 (CDH4) is both significantly DM and expressed during the development of embryonic mice [56]. Finally, HAS1 and Wingless/Integrated 5B (WNT5B) contribute to CAVD [57,58], and SLC16A3 to AS [59].

The network constructed upon the genes linked to DM promoters identified hub proteins known to be involved in different aortic and mitral valve mechanisms. UBC and UBL4A, the two most connected hub genes of the network, belong to the ubiquitin family, whose members play a role in BAVs as well as in atherosclerosis [60,61]. The network’s first submodule was additionally entirely centered around UBC further highlighting the importance of the ubiquitin system, which is key to performing protein QC in the heart [62]. SMAD3, the second most connected hub node, plays an important role in cardiogenesis [63], and is linked to thoracic aortic aneurysm and dissection [64]. RXRA, a member of the RA signaling pathway and the network’s third hub node, is linked to OFT and AV canal malformations, which influence proper aortic and mitral valve development [65]. Finally, the hub node SH3KBP1 is implicated in CAVD [66]. The second submodule of the network contained only two hub nodes APOA5 and BRCA2. Dyslipidemia linked to Lipoprotein a (LPA)-associated APOA5 has been detected in AS, with the reduction of LPA levels via PSCK9 inhibitors constituting promising therapeutic avenues for AS treatment [67–69]. BRCA2, has not been associated with valvular mechanisms in the literature and thus should be further investigated.

Detected pathways were relevant to valvular development and disease (**Table 4**). For example, TGFB- and NOTCH signaling pathways activate EMT by downregulating VE-Cadherin [3,70], which decreases cell adhesion of the transforming endocardial cells enabling them to break away from the endocardium and to migrate into the cardiac jelly, where they can transform into mesenchyme cells creating cushions that expand and fuse to ultimately form cardiac valves [3]. TGFB signaling is also implicated in pathological ECM remodeling [71], MVP [72], AS [73], and NOTCH pathways in BAV and AS [70]. FGF signaling further promotes OFT myocardial cell invasion to the cardiac cushion during EMT, with its disruption leading to malformed OFT valves in mice [74]. Canonical WNT- and Cadherin signaling add to cushion development and remodeling during EMT [3,75], with WNT pathways being additionally implicated in valve stratification and well as in the patterning of the heart forming field [3]. Similar to WNT- and Cadherin-, the PDGF signaling pathway is also involved in cardiogenesis, particularly in the formation of the primordial heart tube [76]. Both the VEGF- and Integrin signaling pathways contribute to post-EMT maturation, in that the former establishes an equilibrium between proliferation and differentiation of cells in the cushion [3], and the latter enables ECM remodeling through the generation of a mechano-transducing network that connects the cells to the ECM providing a link that relays external metabolic and hemodynamic factors [77,78]. HIF-1 signaling is involved in pathological ECM remodeling associated with RMV and MMV disease [79]. Aberrant angiogenesis and IL-17 signaling are also implicated in RMV and infective endocarditis (IE), respectively [80,81]. NF-κB-, TNF-, osteoclast differentiation-, Endothelin and Apoptotic pathways are involved in AS [82–86], with the Endothelin pathway additionally being implicated in the regulation of VICs by VECs [87]. Finally, the detected PPAR signaling pathway is linked to lipid metabolism and is enriched among other lipid-related genes in the second mitral valve-specific subnetwork [88]. The enrichment of PPAR signaling, the uniform increased methylation of the subnetwork’s constituent genes in mitral compared to aortic valves and APOA5 being a hub node, indicate that this subnetwork exhibits major methylation alterations related to the metabolism of lipids. The regulation and expression of lipid-related genes need to be further dissected as it has been reported in a previous study that increased fatty infiltration of valves is observed in MVP [89].

Our analysis provided a comprehensive catalogue of genes and pathways that are differentially regulated between aortic and mitral valves, establishing the basis for upcoming whole-genome bisulfite sequencing (WGBS) studies to additionally interrogate methylation of gene-bodies and other non-promoter regions. Additional insights can be obtained from histone-modification and RNA-based experiments as well as the interaction of such epigenetic mechanisms with DNA methylation. Functional validation of genes and pathways of interest on the transcriptomic and proteomic level will confirm candidate DM biomarkers, which can serve as potential drug targets. Additionally, the same analysis on the VIC level will be done to confirm cell-type specific signals that might have been affected by the tissue’s intrinsic cell heterogeneity. As mentioned above, valves have a unique trilamellar structure housing VICs in a complex consisting of an ECM covered by VECs, all of which contribute to the detected differences. Such analysis will provide novel mechanistic insights into the distinct roles of the individual components.

Limitations – In this study, due to the scarcity of human non-diseased donor heart valves, a relatively small number of valves was examined (n = 12) and two of the four heart valves were studied. Further validation is required to evaluate the clinical significance of the methylation markers identified. An enhancement of the methodology used in this manuscript will be utilized in future studies for example by performing WGBS.

## 5. Conclusion

To conclude, this is the first study that explores the genome-wide DNA methylation landscape characterizing human non-diseased aortic and mitral valves. By investigating genes that are linked to DM promoters and their associated pathways, we discovered that the cells as well as the ECM of the aortic and mitral valve have different methylation signatures. The detected pathways included TGFB-, NOTCH-, FGF-, WNT-, Cadherin- and VEGF signaling pathways associated with EMT, Integrin- and HIF-1 signaling linked to ECM remodeling and NF-κB-, TNF-, osteoclast differentiation, Endothelin- and IL-17 signaling observed in aortic and mitral valve disease. Especially with the increasing incidence and prevalence of valve disease worldwide due to the world’s increasing population age in developed and the failure to address RHVD in low and middle-income countries (LMICs) [6], it is very important to acquire a better understanding of the genetic and epigenetic make-up of cardiac valves and how they are influenced by local conditions, environmental factors and ethnicities as this will affect the development of preventative and therapeutic strategies.

## Data Availability

The data was deposited in the European Genome-phenome Archive (EGA) and is accessible through the accession number EGAD00001006303.

https://www.ebi.ac.uk/ega/studies/EGAS00001004559

## 6. Supplementary Materials

Supplementary materials can be found online.

## 7. Author Contributions

Conceptualization, Najma Latif, Yasmine Aguib and Magdi Yacoub; Data curation, Sarah Halawa; Formal analysis, Sarah Halawa; Funding acquisition, Yasmine Aguib and Magdi Yacoub; Investigation, Sarah Halawa, Najma Latif, Yuan-Tsan Tseng and Adrian H. Chester; Methodology, Sarah Halawa, Najma Latif, Ahmed Moustafa, Yasmine Aguib and Magdi Yacoub; Project administration, Najma Latif and Yasmine Aguib; Resources, Najma Latif, Ayman M. Ibrahim, Adrian H. Chester and Yasmine Aguib; Software, Sarah Halawa; Supervision, Ahmed Moustafa, Yasmine Aguib and Magdi Yacoub; Validation, Sarah Halawa; Visualization, Sarah Halawa, Ahmed Moustafa and Yasmine Aguib; Writing – original draft, Sarah Halawa; Writing – review & editing, Najma Latif, Ahmed Moustafa, Yasmine Aguib and Magdi Yacoub.

## 9. Acknowledgments

We kindly acknowledge Engineer Yehia Raef (Aswan Heart Centre) for supporting this work with scientific data management. We also thank Professor John Chambers and his team for hosting part of Sarah Halawa’s training.

